# Optimizing and Evaluating Robustness of AI for Brain Metastasis Detection and Segmentation via Loss Functions and Multi-dataset Training

**DOI:** 10.1101/2025.08.22.25334255

**Authors:** Yiding Han, Piyush Pathak, Omar Awad, Abdallah S. R Mohamed, Vincent Ugarte, Boran Zhou, Daniel Allen Hamstra, Alfredo Enrique Echeverria, Hasan Al Mekdash, Zaid Ali Siddiqui, Baozhou Sun

**Affiliations:** Baylor College of Medicine

## Abstract

**Purpose:** Accurate detection and segmentation of brain metastases (BM) from MRI are critical for the appropriate management of cancer patients. This study investigates strategies to enhance the robustness of artificial intelligence (AI)-based BM detection and segmentation models.

**Method:** A DeepMedic-based network with a loss function, tunable with a sensitivity/specificity tradeoff weighting factor α- was trained on T1 post-contrast MRI datasets from two institutions (514 patients, 4520 lesions). Robustness was evaluated on an external dataset from a third institution dataset (91 patients, 397 lesions), featuring ground truth annotations from two physicians. We investigated the impact of loss function weighting factor, α and training dataset combinations. Detection performance (sensitivity, precision, F1 score) and segmentation accuracy (Dice similarity, and 95% Hausdorff distance (HD95)) were evaluated using one physician’s contours as the reference standard. The optimal AI model was then directly compared to the performance of the second physician.

**Results:** Varying α demonstrated a trade-off between sensitivity (higher α) and precision (lower α), with α=0.5 yielding the best F1 score (0.80 ± 0.04 vs. 0.78 ± 0.04 for α=0.95 and 0.72 ± 0.03 for α=0.99) on the external dataset. The optimally trained model achieved detection performance comparable to the physician (F1: AI=0.83 ± 0.04, Physician=0.83 ± 0.04), but slightly underperformed in segmentation (Dice: 0.79 ± 0.04 vs. AI=0.74 ± 0.03; HD95: 2.8 ± 0.14 mm vs. AI=3.18 ± 0.16 mm, p<0.05).

**Conclusion:** The derived optimal model achieves detection and segmentation performance comparable to an expert physician in a parallel comparison.

## Introduction

Each year, approximately 100,000 – 240,000 new patients develop brain metastases (BM) [1, 2]. These pose a significant challenge to survival and quality of life for cancer patients [3]. Stereotactic radiosurgery (SRS), which delivers a single, high dose of radiation to BM using either multiple cobalt sources or a linear accelerator, has become a widely adopted treatment option for BM[4–6]. Accurate detection and precise contouring of BM are crucial for successful treatment planning in SRS[7, 8].

Traditionally, the detection and contouring of BM is a labor-intensive and time-consuming process, performed manually by radiation oncologists[9, 10]. This manual approach is subject to human error and significant inter-observer variability, particularly for small or low-contrast lesions that may be easily overlooked or challenging to delineate [11]. Although small metastases can be more easily identified in the follow-up scans after they grow larger, treatment at later stages may be less effective and/or associated with more treatment-related toxicity [12]. The introduction of automated metastasis detection and segmentation could significantly improve image evaluation by reducing human error, enhancing efficiency, and improving the tracking of outcomes [13].

Deep learning models have recently shown great potential in medical image analysis, particularly in tasks such as segmentation and detection [14]. While several studies have achieved a per-patient level high sensitivity rate (true positive (TP) rate), often exceeding 80% -- in detecting BM [10, 15, 16], they frequently suffer from high false-positive (FP) rates [17, 18]. The systematic review by B.B. Ozkara et.al [19] provides a comprehensive list of deep learning models for BMs detection and segmentation.

However, most of these models are trained and tested on datasets from a single institution, limiting their applicability to specific data domains and reducing performance reliability when applied across diverse imaging protocols and equipment. Additionally, per-patient metrics, which aggregate results across all lesions in a patient, tend to disproportionately weight patients with fewer lesions. In datasets where single-lesion cases dominate, high per-patient sensitivity and precision can mask poor performance in multi-lesion cases, which are often more clinically challenging.

Another fundamental challenge in developing and evaluating AI for BM is the inherent uncertainty in defining ‘ground truth’ from physician annotations [16]. Inter-physician variability in contouring, particularly for challenging lesions, is well-documented [11]. For example, Ziyaee et.al’s [15] study on AI segmentation performance found that 43% of the lesions graded as false positives were actual tumors, based on secondary review or follow up. This underscores the limitations of evaluating AI solely against a single physician’s contours as the definitive standard. To address these limitations and improve the robustness of AI models for BM detection and segmentation, this study aims to systematically evaluate two key strategies: optimizing the loss function and leveraging diverse multi-institutional training datasets. We determine how different configurations impact performance, particularly on lesion-level metrics and as a function of tumor volume, to identify practical approaches applicable across varying clinical settings. Furthermore, rather than assuming a single physician’s segmentation represents absolute ground truth, we adopt a more rigorous evaluation approach: a parallel comparison between the developed AI model and an experienced radiation oncologist, using a separate physician’s contours as a standard reference. This framework provides a more objective assessment of AI performance relative to clinical practice, explicitly accounting for inter-physician variability and offering a fairer benchmark for clinical integration.

Thus, this study contributes:(1) an investigation of the impact of loss function weighting and diverse training data compositions on AI model robustness for BM detection and segmentation, particularly evaluating performance as a function of tumor volume on an external, multi-physician dataset; (2) strategies for building more robust models, including dataset combination approaches tailored to lesion size; and (3) p a practical framework for evaluating AI against human performance by implementing a parallel comparison approach that accounts for inter-observer variability inherent in clinical annotation.

## Method and Materials

### Patients

This study utilized T1 post-contrast MRI brain imaging datasets from three institutions to train and evaluate AI models for brain metastasis detection and segmentation.

I. **UCSF BMSR Dataset**: The primary dataset for training was the University of California San Francisco Brain Metastasis Stereotactic Radiosurgery (UCSF BMSR) dataset [20]. It comprises 412 patients (238 female and 174 male) with a total of 461 imaging series, including 314 pretreatment cases and 147 follow-up cases. BM segmentations were provided by attending physicians or neuroradiology fellows, serving as the reference standard and internal validation. MRIs were acquired on a 1.5-T GE SingaHDxt scanner with TR = 8.8 ms, TE = 3.3 ms, flip angle = 11°.
II. **Our Institution Dataset**: The second training dataset was collected at in our own institution and includes 53 patients (28 female, 25 male) with imaging acquired between January 1, 2018, and March 7, 2024. Attending radiation oncologists provided the reference standard BM segmentations as part of routine clinical care for SRS delivery. MRIs were acquired with Sagittal fat-saturated 3D T1-weighted variable-flip-angle turbo spin-echo (SPACE) acquisition, reformatted into axial 1 mm isotropic voxels (Siemens).
III. **The Cancer Imaging Archive (TCIA) Dataset**: The third dataset, published on The Cancer Imaging Archive, as “GammaKnife-Hippocampal” (TCIA-GK-Hippo) [21], consists of pre-treatment imaging from 91 patients (61 female and 30 male) undergoing Gamma Knife radiosurgery. This dataset was then annotated by independent segmentations provided by two senior trainees (PGY-5 or higher) radiation oncologists, O.A. and P.P., for all 91 MRI series for the purpose of this study. In this study P.P.’s segmentations were designated as the reference standard (‘ground truth) for evaluating model performance, while O.A.’s segmentations were utilized to represent the performance of an expert physician in the parallel comparison. Both physicians complete the segmentation on a Brainlab workstation (Brainlab AG, Munich, Germany). MRIs were acquired with 3-D T1-weighted spoiled gradient-echo (FLASH) sequence, post-contrast, on a 1.5 T Siemens system: TR = 11 ms, TE = 3.2 ms, flip angle = 20°.

The use of institutional data is covered by an IRB-approved research plan. Clinical and demographic information was not collected for this study.

### Data preprocessing

Images were first converted from Digital Imaging and Communications in Medicine format into Neuroimaging Informatics Technology Initiative (NIfTI) format. To protect personal health information, all images were skull-stripped [22]. The images underwent intensity normalization and were resized to 1.0 × 1.0 × 1.5mm spacing.

### Model architecture and training parameters

We utilized the DeepMedic network architecture [24], a multi-scale 3D convolutional neural network, as the baseline method for this study. The network processes input segments for the main pathway consisting of 37^3^ voxels, using nine convolutional layers with 3^3^ kernels without padding, resulting in a receptive field size of 19^3^. The effective segment size in the network output is also 19^3^. Two additional parallel convolutional pathways with lower resolutions (scaled by factors of 3 and 5) were included to capture context at different scales.

A significant challenge in this work is the class imbalance, as normal tissue voxels vastly outnumber tumor voxels. To address this, the training process included volume segmentation (calculating the volume of background vs. target) with sampling designed to maintain class balance in the samples presented to the model.

The initial learning rate was set to 0.001, and optimization was performed using the RMSProp optimizer combined with Nesterov momentum [23] with the moment value m = 0.6, *ρ* = 0.9, and *ϵ* = 10^−4^.

### Multi-dataset training

To evaluate model robustness and generalizability, we trained models using three different training dataset compositions:

1. **Main Training Dataset**: This dataset consisted solely of UCSF data only. Of the 412 patients available, 308 were used for training and 104 for internal validation. This dataset is labeled as “main training” in Figure 1.
2. **Heterogeneous Training Dataset**: This dataset combines UCSF data (308 patients) and our internal data (53 patients, 92 lesions). Our internally collected data under different clinical conditions and representing a smaller cohort, was included to introduce variability, thus labeled “Heterogeneous Training”.
3. **TCIA Few-Shot Training Dataset**: This dataset augmented the “Heterogeneous Training” dataset (UCSF +our internal data) with a small subset of TCIA data. Specifically, 10 randomly selected pre-treatment patients from the external testing dataset labeled as “TCIA” in Figure 1 were added. This approach simulated a scenario where limited data from a target institution is available for fine-tuning or adapting a model trained on external data. This small subset constituted less than 5% of the total training data volume. Each training dataset reflects distinct aspects of clinical variability, providing insights into how different data sources impact model robustness and generalizability.

**Figure 1.**
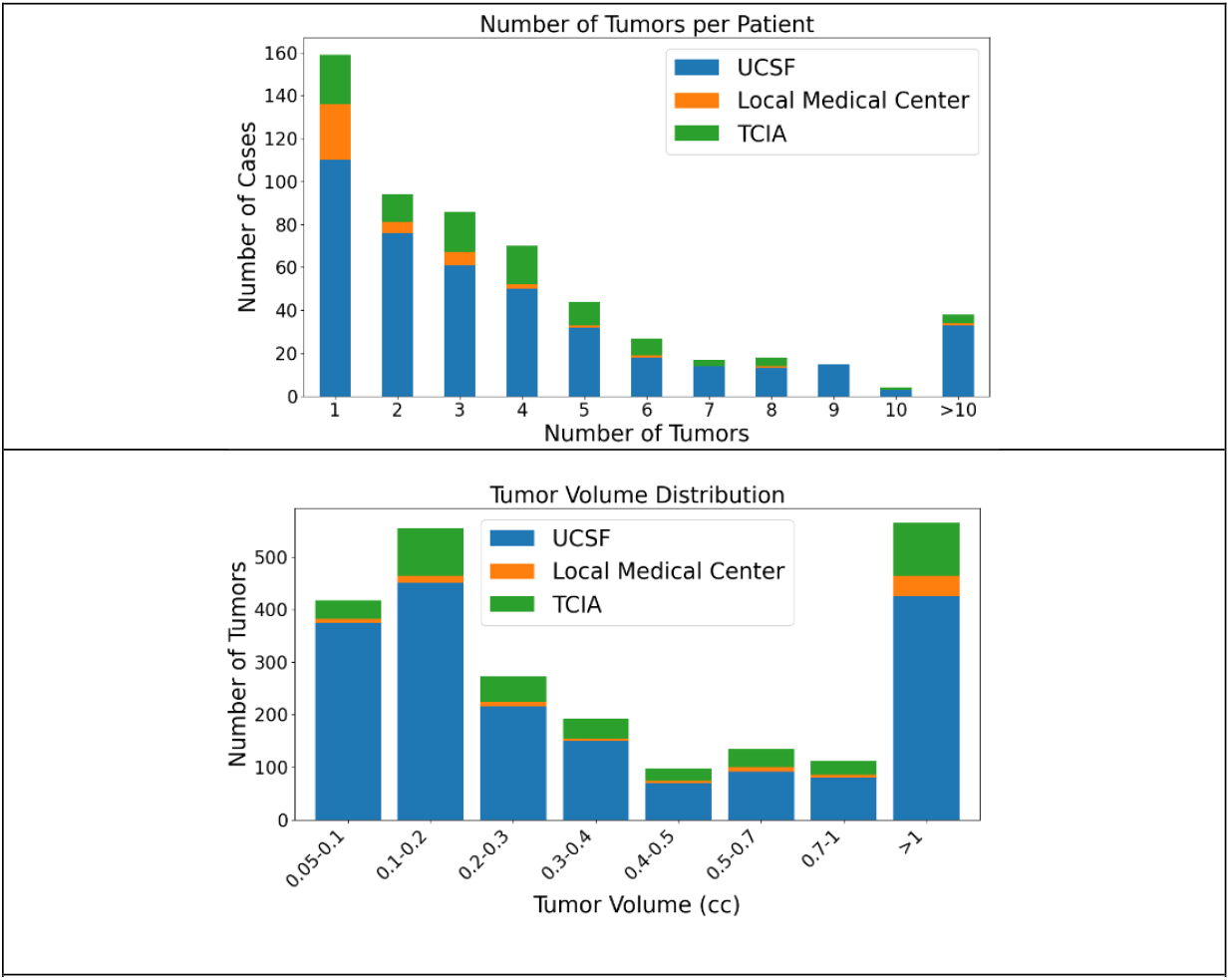
The metastasis statistics for the datasets used in this study

### The test dataset in all experiments was a hold-out cohort from the TCIA that was not used for model training

#### Loss function

We employed a custom loss function that combined a volume-level sensitivity-specificity (VSS) loss function designed to enhance BM detection performance. The loss function combines the VSS loss (*l*_*vss*_)and binary cross-entropy loss (BCE, *l*_*BCE*_)

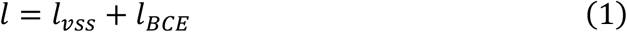

The binary cross-entropy loss [24, 25] is defined as:

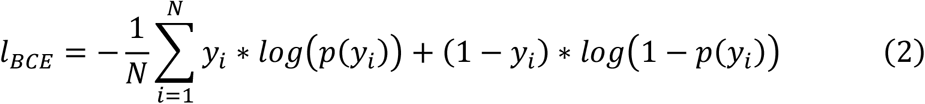

- *y*_*i*_ is the segmentation label of the i-th volume.
- *p*(*y*_*i*_) is the model prediction of the i-th volume.

The VSS loss, designed to balance volume-level detection performance, is defined as:

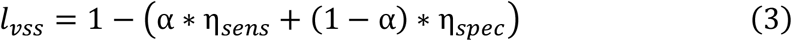

Here, *η*_*sens*_ is Volume-level sensitivity, defined as:

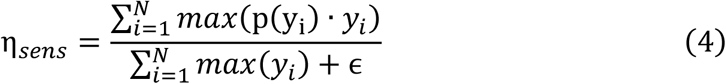

And *η*_*spec*_ is Volume-level specificity, defined as:

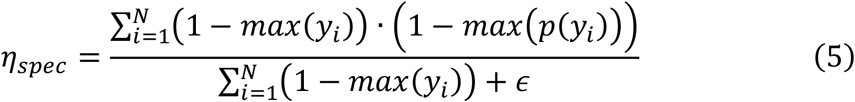

ϵ is a small constant added to avoid division by zero, and α is a weight parameter that balances the importance of sensitivity (η_*sens*_) and specificity (η_*spec*_). Notably, emphasizing volume-level specificity (lower α values) is intended to reduce false positives at the volume level, thereby improving detection precision at the lesion level.

In this study, we trained models in three different weight factors for each training dataset: α = 0.5, 0.95, 0.99. This resulted in a total of nine distinct models (3 α values × 3 training datasets). Experiments varying α exclusively utilized the UCSF dataset for training.

#### Evaluation metrics

For clinical applications, both accurate detecting and segmenting BM are critical. Our evaluation focuses on metastasis-level metrics, calculated after applying a probability threshold to the network output and performing connected component analysis to identify individual lesions. detection performance is measured using:

- **Sensitivity:** TP / (TP + FN) - the proportion of true metastases correctly detected.
- **Precision:** TP / (TP + FP) - the proportion of detected lesions that are true metastases.

True Positives (TP), False Positives (FP), and False Negatives (FN) are determined at the metastasis level by comparing detected lesions to the reference contours.

Segmentation accuracy is evaluated using two standard metrics for comparing predicted segmentations (A) with reference segmentations (B):

- **Dice Similarity Coefficient (DSC):** DSC = 2|A ∩ B| / (|A| + |B|) [29]. This measures the spatial overlap between the predicted and reference contours.
- **95th Percentile Hausdorff Distance (HD95):** The Hausdorff Distance measures the maximum distance between points in two sets. HD95 [30] is a more robust variant that calculates the 95th percentile of these distances, reducing sensitivity to outliers and providing an indication of spatial discrepancy.

All models were initially evaluated on the external TCIA dataset using P.P.’s contours as the reference standard. Performance metrics (sensitivity, precision, F1 score for detection; Dice, HD95 for segmentation) were analyzed as a function of tumor volume.

Based on these evaluations across different loss function weightings and training datasets, an “optimal model strategy” was identified (details presented in the Results section). This strategy involved combining predictions from models trained on different dataset compositions, tailored to tumor volume. The performance of this “optimal model strategy” was then directly compared to the performance of the second physician (O.A.) on the same TCIA dataset, again using P.P.’s contours as the reference standard. Differences in performance were assessed using the Wilcoxon signed-rank test, a non-parametric method for paired data, with a significance threshold = 0.05. This parallel comparison aimed to provide a clinically relevant benchmark for the AI model’s capabilities relative to an experienced human expert, explicitly acknowledging inter-observer variability by evaluating both against a common reference.

## Results

The characteristics of the datasets used in this study are summarized in **Figure 1**. Across all datasets, the median number of tumors per patient was 3 with Interquartile range (IQR) = 4 (UCSF:3 with IQR = 4, BCM: 1 with IQR = 2, TCIA: 3 with IQR = 3) with a median tumor volume of 0.258 cc with IQR = 0.76 cc (UCSF:0.234 cc with IQR = 0.723 cc, BCM: 0.657 cc with IQR = 0.412 cc, TCIA: 0.363 cc with IQR = 0.895 cc). 5%, 0%, and 2% of the tumors were tumor cavities in each of the datasets, respectively.

### Effect of Loss Function Weight Factor (α)

Figure 2 illustrates the detection sensitivity (upper left), precision (upper right), and F1 score (lower) as functions of tumor volume for models trained with α = 0.5, 0.95, and 0.99. The shaded bands represent statistical uncertainty.

**Figure 2:**
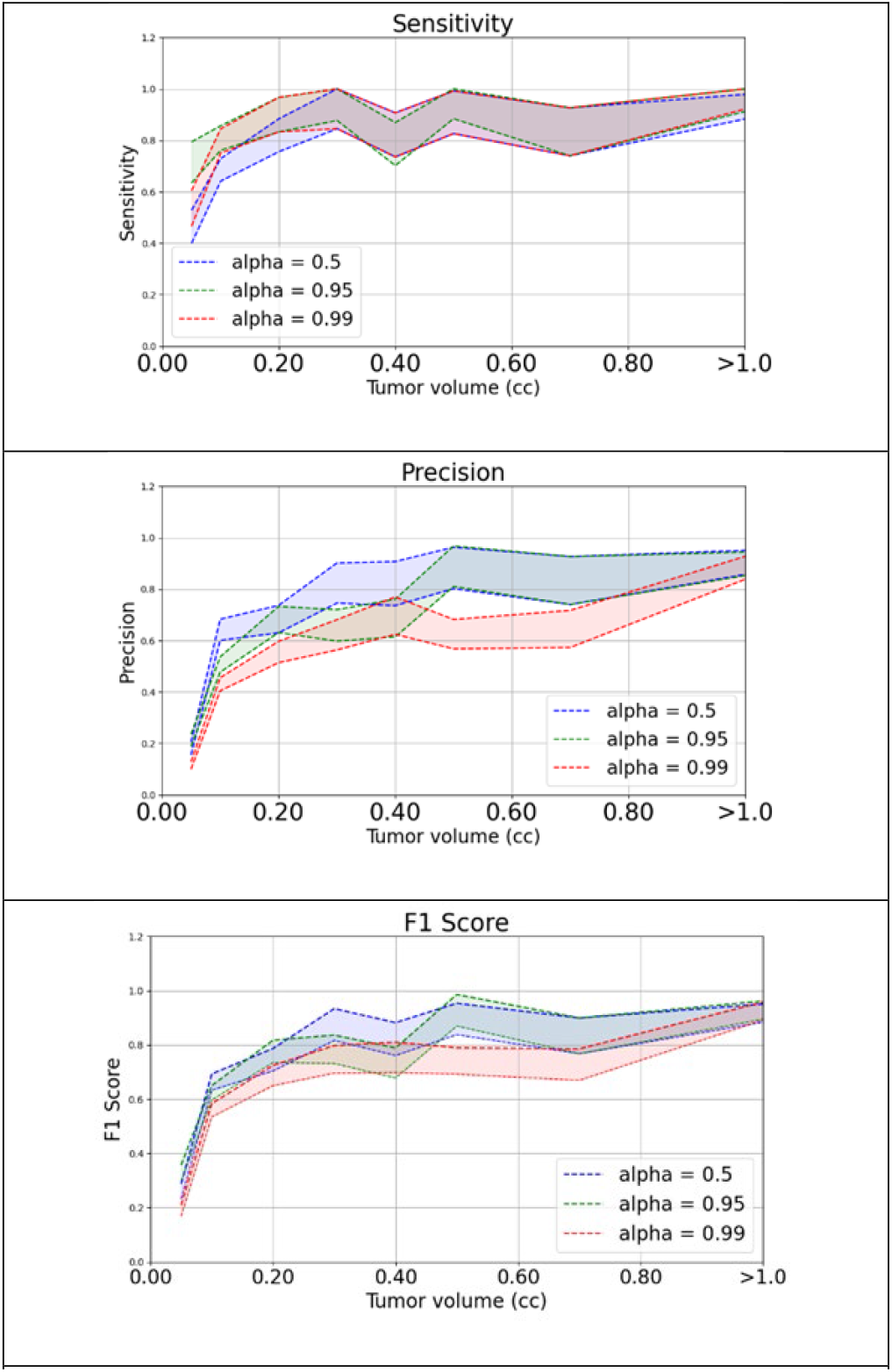
the detection sensitivity (upper), precision (middle), and F1 score (lower) for models with different weight factors, α=0.5 (blue), 0.95 (green), 0.99 (red), in the loss function

The results show a clear trade-off: higher α values (emphasizing sensitivity) lead to improved detection sensitivity, particularly for smaller tumor volumes (<0.4 cc), but result in lower precision due to an increased number of false positive detections. Conversely, lower α values (emphasizing specificity/precision) yield higher precision but lower sensitivity, especially for small lesions. This effect is visually demonstrated in Figure 3, which shows example cases highlighting how higher α improves detection of small lesions (left panel) but can increase false positives (right panel).

**Figure 3.**
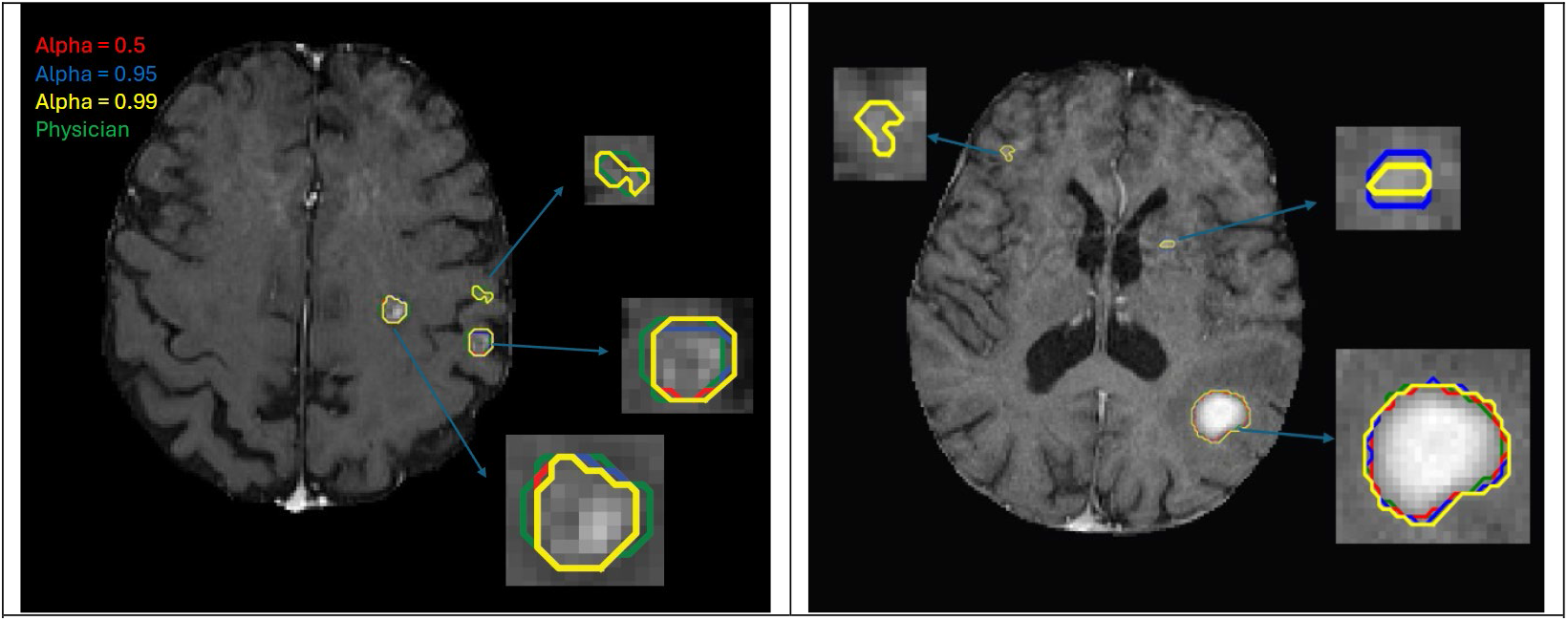
Two examples of the BMs detection sensitivity (left panel) and precision (right panel) to illustrate the weight factor effect with α = 0.5 (red), 0.95 (blue), 0.99 (yellow), and physician (green) as reference.

Table 1 summarizes the average F1 scores across all tumor volumes for each α value. The model trained with **α = 0.5 achieved the best overall average F1 performance (0.80 ± 0.04)** compared to α = 0.95 (0.78 ± 0.04) and α = 0.99 (0.72 ± 0.03). Based on these results, α = 0.5 was selected as the optimal weight factor for subsequent experiments exploring the effect of training datasets.

**Table 1.**
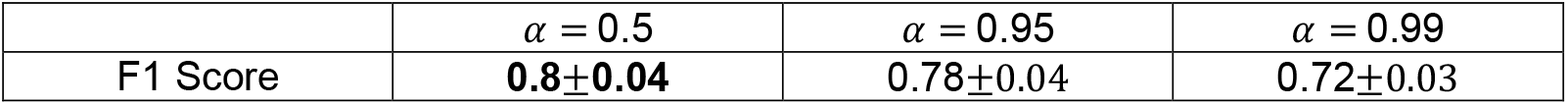
Average F1 score for tumor detection with different weight factors.

| | $\alpha = 0.5$ | $\alpha = 0.95$ | $\alpha = 0.99$ |
| --- | --- | --- | --- |
| F1 Score | **$0.8 \pm 0.04$** | $0.78 \pm 0.04$ | $0.72 \pm 0.03$ |

### Effect of Multi-institutional training datasets (with α = 0.5)

Figure 4 displays the sensitivity (top), precision (middle), and F1 score (bottom) as functions of tumor volume for models trained on the UCSF dataset alone, UCSF+BCM(“Heterogeneous Training”), and UCSF+Our Institution+TCIA Few-Shot (“Few-shot Training”). The shaded bands represent statistical uncertainty. All evaluations were performed on the external TCIA dataset.

**Figure 4:**
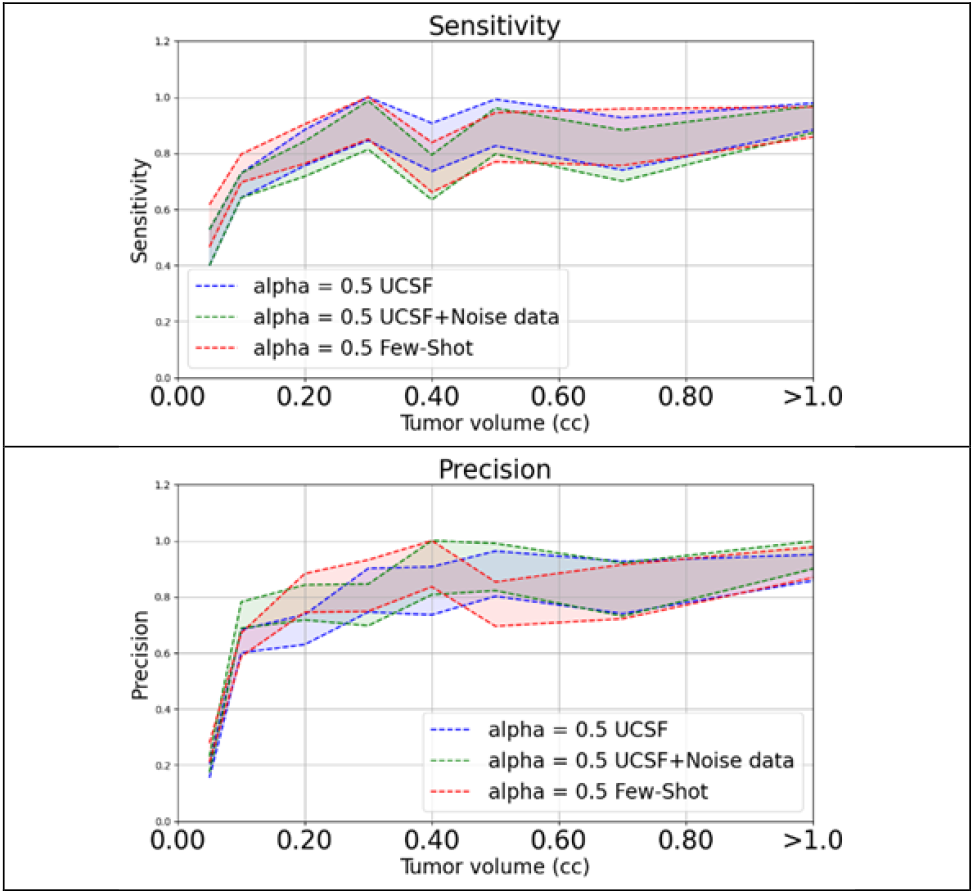
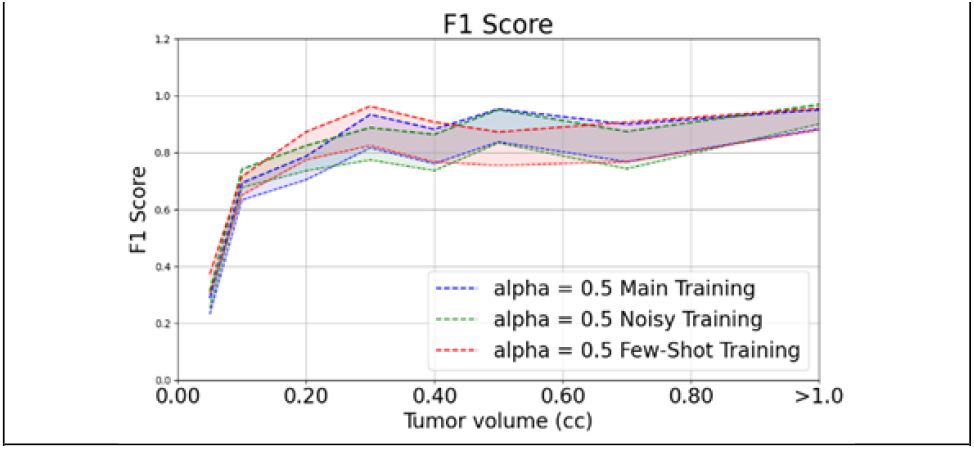
the detection sensitivity (upper), precision (middle), and F1 score (lower) for models with different training datasets, Main Training: UCSF only (blue), Heterogeneous Training: UCSF+Our Institution (green), Few-shot Training: UCSF+Our Institution+TCIA Few-shot (red).

Figure 5 illustrates two examples to show performance variation between the different training datasets strategies.

**Figure 5:**
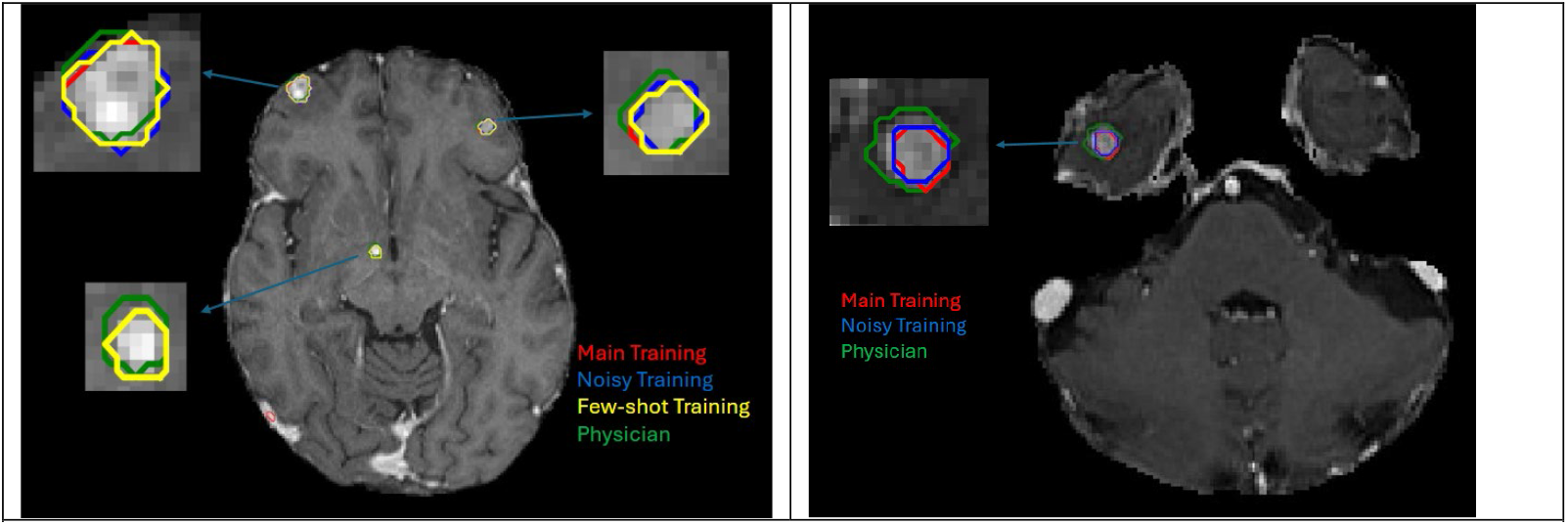
Two examples of the BMs detection sensitivity (left panel) and precision (right panel) to illustrate the various training dataset *effects* with Main Training (red), Heterogeneous Training (blue), Few-shot Training (yellow), and physician (green) as reference.

Table 2 summarizes the average F1 scores specifically for tumor volumes < 0.4 cc and > 0.4 cc across the different training datasets (all using α=0.5).

**Table 2.** The F1 score for different training datasets on different tumor volumes +/- standard deviation.

| | Tumor volume $< 0.4$ cc | Tumor volume $> 0.4$ cc |
| --- | --- | --- |
| Main Training | $0.69 \pm 0.06$ | $0.89 \pm 0.07$ |
| Heterogeneous Training | $0.71 \pm 0.07$ | **$0.90 \pm 0.07$** |
| Few-shot Training | **$0.72 \pm 0.07$** | $0.87 \pm 0.07$ |

These results indicate that no single training dataset composition is optimal for all tumor sizes. The Few-shot training improves performance for small tumors, while the Heterogeneous training improves performance for larger tumors.

### Optimal Model Strategy and Physician Comparison

Although the different dataset compositions showed no statistically different results in training strategy based on dataset composition, we tested results on, which showed that the Few-shot trained model was best for tumors < 0.4 cc and the Heterogeneous trained model was best for tumors > 0.4 cc, we developed an **Optimal Model Strategy**. This strategy involves combining the strengths of the models trained on different datasets by using the predictions from the **Few-shot trained model (α=0.5)** for detected lesions with a volume **less than 0.4 cc**, and the predictions from the **Heterogeneous trained model (α=0.5)** for detected lesions with a volume **greater than 0.4 cc**.

We then performed a parallel comparison between this Optimal Model Strategy and the performance of the **second physician (O.A.)** on the external TCIA dataset. For this comparison, the segmentations provided by the **first physician (P.P.)** were used as the reference standard (‘ground truth’).

Figure 6 compares the detection performance (sensitivity, precision, and F1 score) of the Optimal Model Strategy (blue) and the second physician (red) as functions of tumor volume on the TCIA dataset. The shaded bands represent statistical uncertainty.

**Figure 6:**
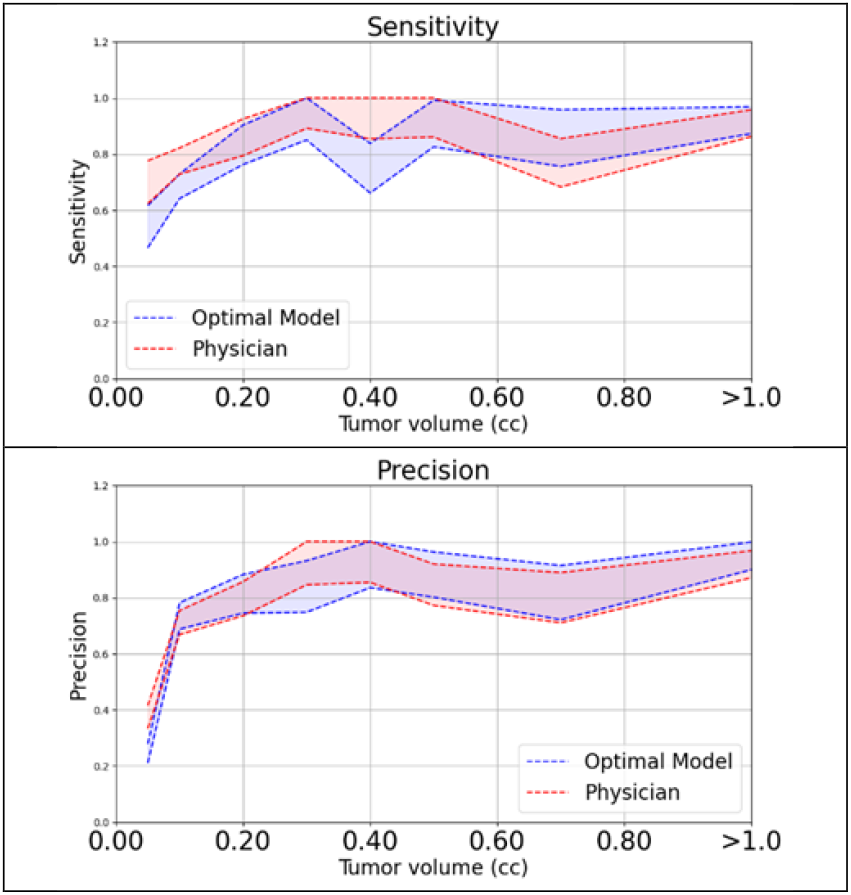
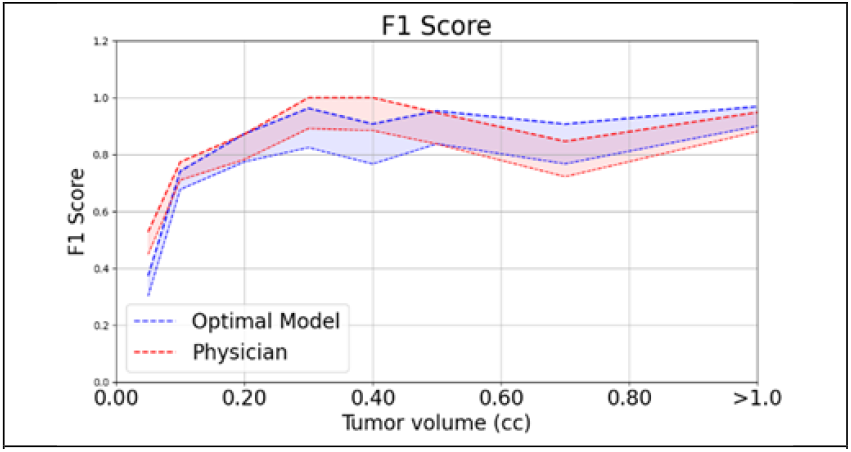
the detection sensitivity (upper), precision (middle), and F1 score (lower) for the optimal model (blue) and physician (red), the shaded bands represent the statistical uncertainty.

### Segmentation

Figure 7 compares the segmentation performance (Dice and HD95) between the Optimal Model Strategy (blue) and the second physician (red) on the same dataset, also as functions of tumor volume.

**Figure 7:**
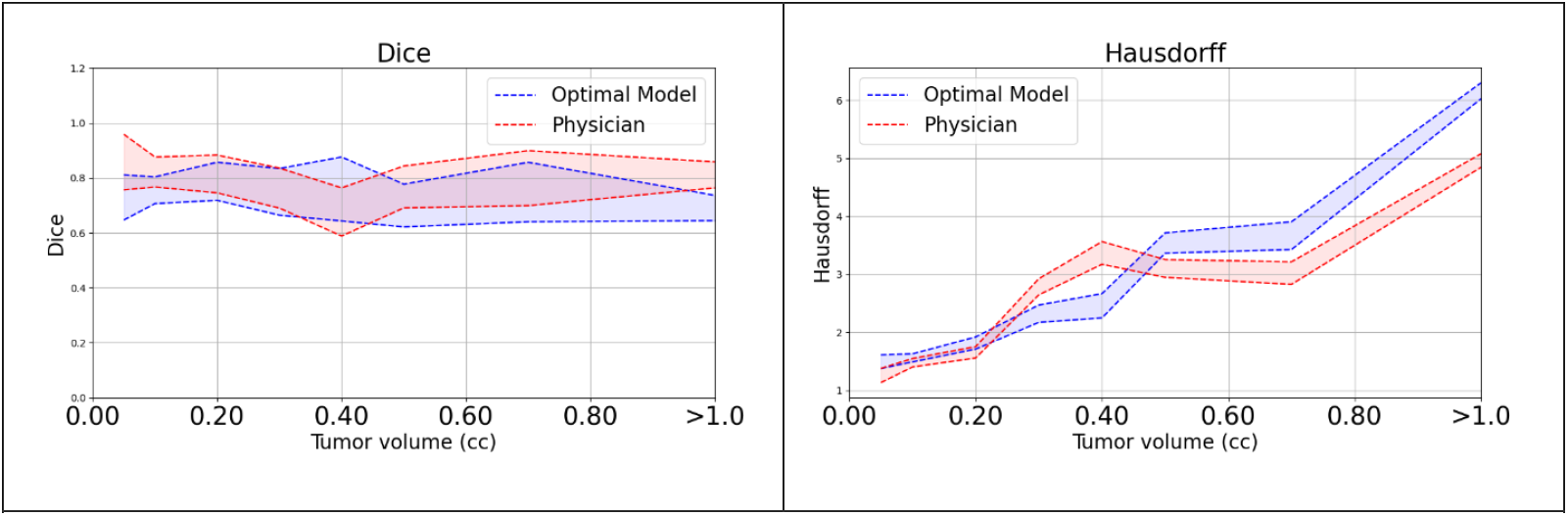
The segmentation Dice score (left) and HD95 (right) for the optimal model (blue) and physician (red), the shaded bands represent the statistical uncertainty.

Table 3 summarizes the average detection and segmentation performance metrics across all tumor volumes for the Optimal Model Strategy and the second physician (O.A.), using the first physician (P.P.) as the reference.

**Table 3.**
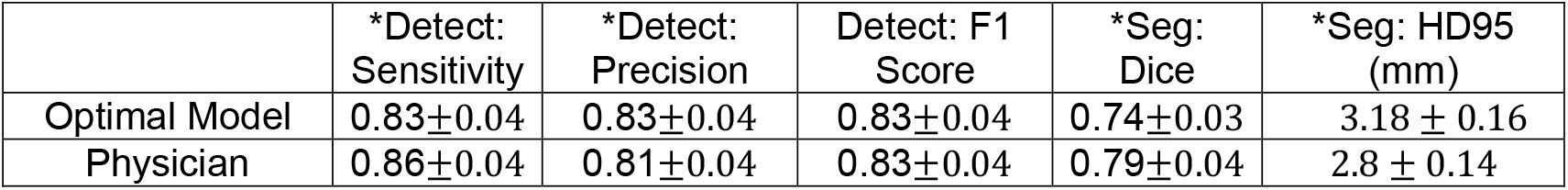
Average detection and segmentation performance of the optimal model and the second physician. * indicates p-value <0.05 for between-group comparison

|  | *Detect:<br>Sensitivity | *Detect:<br>Precision | Detect: F1<br>Score | *Seg:<br>Dice | *Seg: HD95<br>(mm) |
| --- | --- | --- | --- | --- | --- |
| Optimal Model | $0.83 \pm 0.04$ | $0.83 \pm 0.04$ | $0.83 \pm 0.04$ | $0.74 \pm 0.03$ | $3.18 \pm 0.16$ |
| Physician | $0.86 \pm 0.04$ | $0.81 \pm 0.04$ | $0.83 \pm 0.04$ | $0.79 \pm 0.04$ | $2.8 \pm 0.14$ |

The results in Table 3 show that the Optimal Model Strategy achieves average detection performance statistically equivalent to the second physician, with identical average F1 scores. However, the second physician demonstrates higher segmentation accuracy based on Dice and HD95.

Figure 8 provides qualitative examples of BM segmentation from the TCIA dataset, illustrating varying levels of agreement among the first physician (P.P., reference), the second physician (O.A., comparison), and the Optimal Model Strategy. The upper panel shows a case with strong agreement among all three, while the lower panel highlights a case with notable discrepancies, particularly in the spatial extent and shape of the segmentation contours, demonstrating inter-observer variability and differences in AI performance relative to each physician.

**Figure 8.**
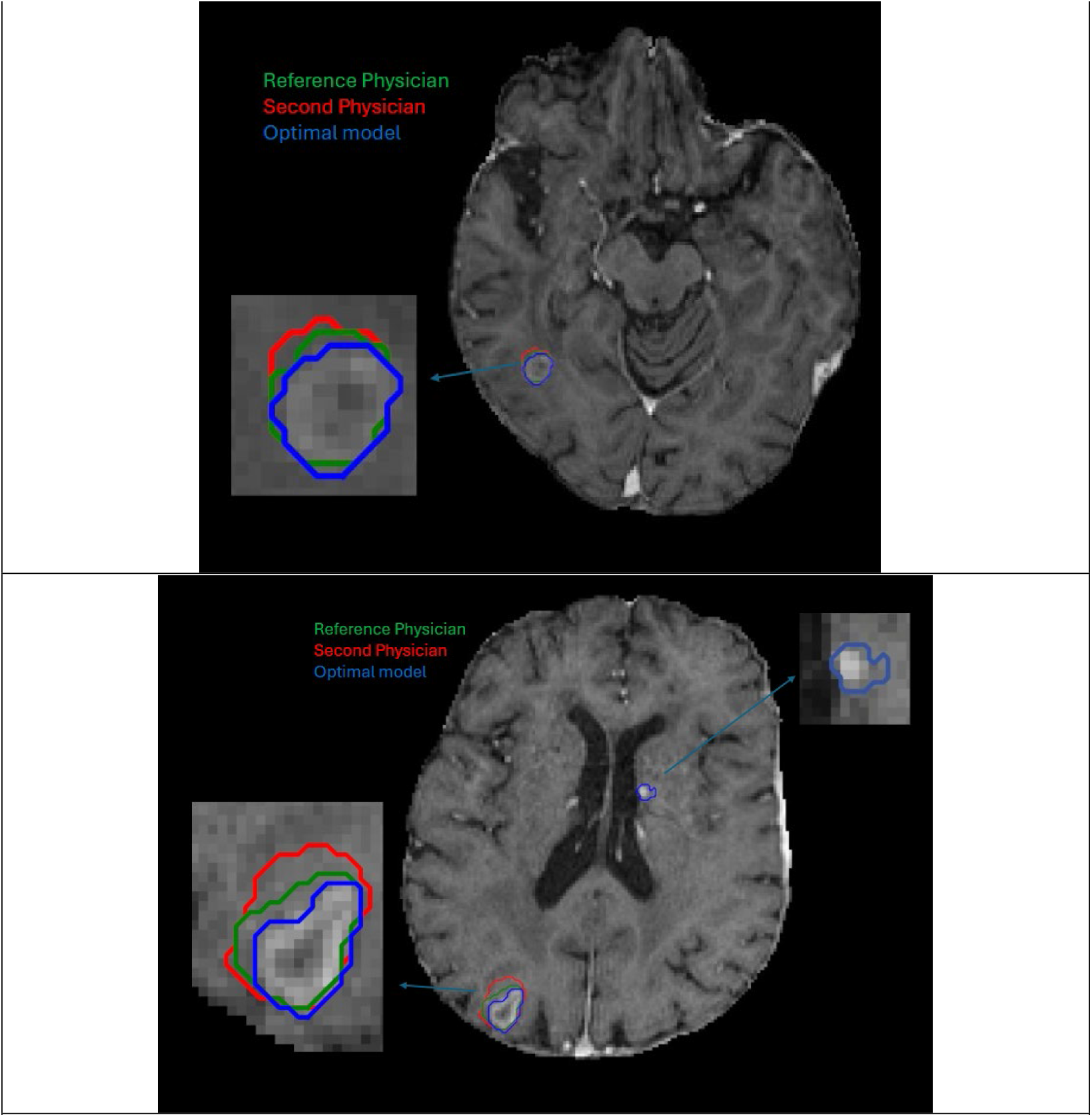
Two examples of BM segmentation illustrate varying levels of agreement: the upper panel demonstrates strong agreement, while the lower panel highlights poor agreement. The comparison includes the optimal model (blue), the second physician (red), and the first physician (green) as the reference.

## Discussion

Our study demonstrates a strong dependence of AI detection performance on tumor volume, consistent with clinical intuition and previous work [26] (Figures 3, 5). Smaller tumors, due to lower signal contrast and limited representation across few image slices, pose a significant challenge for AI models. This difficulty is mirrored in human performance; as illustrated in Figure 6, even experienced physicians exhibit greater variability and reduced performance for detecting small tumors compared to larger ones. This inter-expert variability in identifying and delineating small metastases presents a major challenge in establishing a reliable ‘ground truth’ for AI training and evaluation.

Regarding the impact of the loss function weight (α), our findings (Figure 2) demonstrate the expected trade-off: higher α (greater sensitivity emphasis) improves sensitivity, particularly for small lesions (Figure 3, left), but reduces precision due to increased false positives (Figure 3, right). Conversely, lower α (greater specificity emphasis) enhances precision. Our VSS loss formulation specifically includes a volume-level specificity term (η_*spec*_), and assigning a higher weight to this term (lower α) is intended to penalize false positive detections more heavily at the volume level, thereby improving detection precision at the lesion level. Huang, Y., et al. [27] reported higher sensitivity with increased α and lower precision with decreased α. However, their study relied on an internal dataset for both training and testing, limiting insights into generalization. In contrast, our evaluation on an external dataset demonstrates that for α > 0.95, sensitivity plateaus in most cases, and decreases for tumors < 0.1 cc, highlighting generalization limitations at high α value. This divergence underscores the institutional dependence of model behavior and the importance of external validation. Meanwhile, our evaluation revealed that a more balanced α=0.5 yielded the best overall F1 score (Table 1). This suggests that optimizing the loss function to incorporate volume-level specificity and explicitly penalize false positives can improve model robustness and generalizability across different data domains.

Our evaluation of different multi-institutional training datasets (Figure 5, Table 2) highlights their crucial role in enhancing model robustness. Incorporating data from our institution alongside UCSF data (the ‘Heterogeneous Training’ dataset) generally improved performance for larger tumors (>0.4 cc). Furthermore, adding a small few-shot sample (10 patients) from the TCIA dataset to the training data (‘Few-shot Training’) may boost performance for **small tumors (<0.4 cc)**. These findings strongly support a practical strategy for improving AI generalizability in clinical settings: leveraging diverse training data and employing few-shot or fine-tuning approaches with limited local data can enhance performance across different imaging environments, mirroring successful strategies in large language models and foundation models [28, 29].

Based on the volume-dependent performance observed with different training datasets, we constructed an **Optimal Model Strategy** by combining predictions from the few-shot trained model (optimized for small tumors) and the Heterogeneous trained model (optimized for larger tumors), as detailed in the Results section. We then performed a parallel comparison of this AI strategy against the **second physician (O.A.)** on the external TCIA dataset, using the **first physician’s (P.P.)** contours as the reference standard (Table 3, Figures 6, 7). The average detection F1 score of the Optimal Model Strategy (0.83 ± 0.04) was statistically equivalent (P value < 0.05) to that of the second physician (0.83 ± 0.04), suggesting comparable overall detection capability on this external dataset. However, the second physician demonstrated quantitatively superior segmentation performance based on traditional Dice (0.79 vs. 0.74) and HD95 (2.8 mm vs. 3.18 mm) metrics.

It is crucial to interpret these comparisons within the context of inherent inter-observer variability and the lack of absolute ground truth [20]. While the metrics show the physician outperforming the AI in segmentation *relative to the specific reference contours*, Figure 8 illustrates cases where segmentations from the two physicians **also** differ significantly, and the AI’s segmentation might appear visually more plausible or capture the tumor core more accurately in certain instances (e.g., lower panel). This highlights that traditional geometric metrics like Dice and HD95, calculated against a single, potentially variable reference, may not fully capture the clinical utility or biological plausibility of a segmentation. Future work could explore alternative evaluation approaches, such as assessing segmentations based on predicted clinical outcome or leveraging AI’s capability to produce nuanced probability maps, where each voxel is assigned a probability score. This capability, where AI models produce probability maps centered on tumor cores, could potentially allow for a more fine-grained assessment of confidence at the voxel level, offering a more robust evaluation framework less sensitive to the precise boundaries defined by any single annotator and potentially useful for tasks like grading resident contours during training.

Future research directions could further enhance AI performance and clinical integration. Incorporating longitudinal imaging data, comparing current scans to tumor-free baselines, has shown promise [20, 27] but is less practical for new or emergency cases lacking prior imaging. A more widely applicable approach involves integrating AI models with radiomics features [30–32]. Radiomics has been successfully used to characterize tumor properties and differentiate similar-looking entities like radiation necrosis from true progression [33–35]. Combining deep learning detection with radiomic analysis could help refine the classification of potential lesions, potentially improving the precision of high-sensitivity models (like those trained with higher alpha) by reducing false positives, building upon recent work in other domains [36]. Ensemble methods, combining outputs from multiple models trained with different configurations, could also potentially further boost robustness and performance.

## Conclusion

This study demonstrates that optimizing loss function weighting and strategically utilizing diverse multi-institutional training datasets are effective strategies for developing more robust and accurate AI models for brain metastasis detection and segmentation. While the derived optimal AI model achieves detection performance statistically comparable to an expert physician, inherent inter-observer variability in clinical ground truth necessitates careful consideration in both model training and evaluation.

## Data Availability

All data produced in the present study are available upon reasonable request to the authors

